# Postpartum hyperglycemia and associated factors among women attending postnatal clinics in Dar es salaam: a cross-sectional study

**DOI:** 10.1101/2025.10.14.25338023

**Authors:** Marietha Holela, Bruno Sunguya, Kaushik Ramaiya, Amani Kikula

## Abstract

**Background:** The prevalence of Gestational Diabetes Mellitus in Tanzania is raising paralleling its contribution to maternal morbidity and mortality. Studies have reported a prevalence of postpartum hyperglycemia in Africa to be 15% with 48% of the women progressing to T2DM five years postpartum. Carbohydrate dense diet has been correlated with glucose intolerance in most pregnant women. However, evidence is scarce and not systematic on factors associated with post-partum hyperglycemia in Tanzania. This study assessed nutritional and lifestyle factors associated with postpartum hyperglycemia among women attending postnatal clinics in Dar es salaam.

**Methods:** We conducted an analytical, hospital based cross-sectional study in postnatal clinics based in Dar es Salaam, Tanzania. A multistage simple random sampling method was used to determine the three health centers and recruit women attending postnatal clinic from April to June 2025. Structured questionnaires were used to collect data, descriptive statistics was done using mean, proportion and chi-square test. We employed a univariable and multivariable modified poisson regression analysis to determine the association of independent variables with postpartum hyperglycemia. The p-value of <0.05 was considered statistically significant.

**Results:** We enrolled 392 postpartum women, from Sinza health center (HC) (48%), Magomeni HC (28.3%) and Kinyerezi HC (23.7%) with the mean age of 31.52 (±7.54). Postpartum hyperglycemia was prevalent among 18.6% of the women. The prevalence of postpartum hyperglycemia was higher among women with higher BMI (overweight and obese) (APR= 8.1, p<0.001, 11.6, p<0.001) respectively, and those with refined carbohydrate dietary pattern (APR= 1.54, p=0.022), while diet rich in legumes and vegetables was protective against postpartum hyperglycemia (APR= 0.69, p=0.001), after adjusting for potential confounders.

**Conclusion:** About one in five postpartum women had hyperglycemia and it is linked to modifiable nutritional and dietary factors. These include overweight, obesity and consumption of refined carbohydrate, while protein and legumes were protective factors. Attention should be put to prioritizing nutrition education and promotion of healthy lifestyle during peripartum period to ensure optimal glucose control in the postpartum period.

## Introduction

Postpartum hyperglycemia, characterized by elevated blood glucose levels following childbirth, is increasingly recognized as a significant health concern for women globally [1]. The condition is associated with lasting consequences, including the development of type 2 diabetes mellitus (T2DM) and cardiovascular diseases [2]. The prevalence is notably higher among women with a history of gestational diabetes mellitus, with estimates suggesting that 20-70% may develop persistent hyperglycemia in the first ten year after delivery [3]. Globally gestational diabetes mellitus affects approximately 14% of the pregnancies worldwide [4]. Gestational diabetes poses a 7-10-fold increased risk of chronic type two diabetes to the affected women as opposed to normal glycemic women [5]. The disparities in the prevalence of postpartum hyperglycemia are due to lack of follow up and low uptake of screening tests among women after delivery.

In Sub-Saharan Africa, studies indicate that 45 – 48% of women with gestational diabetes mellitus (GDM) develop T2DM within six years postpartum, with 31% exhibiting persistent hyperglycemia 20% prediabetes and 11% T2DM) at 4 – 12 weeks postpartum [6, 7]. Gestational diabetes mellitus is a public health challenge, particularly in resource-limited settings like Sub-Saharan Africa, where healthcare access is constrained [8]. In Tanzania, the prevalence of GDM has risen sharply, from 5.1% in 2014 to 39% in 2020, driven by nutrition, demographic and demographic transitions affecting lifestyle and food choices [9, 10]. This trend parallels the increasing burden of T2DM, affecting 11.9% of Tanzanian adults [11].

Nutritional factors, including high consumption of refined carbohydrates and legumes, contribute to elevated blood glucose levels due to their high glycemic index, increasing GDM risk [12, 13] Maternal obesity, body fat percentage, and mid-upper arm circumference (MUAC) >28 cm are also linked to GDM and T2DM, as they impair pancreatic beta-cell function and insulin secretion [14–16]. Conversely, lifestyle modifications, such as moderate to vigorous physical activity (e.g., 30 minutes daily or 500 metabolic equivalent of task (MET-min/week), enhance insulin sensitivity and glucose regulation, offering protective effects against hyperglycemia [17–19].

Despite the growing burden of GDM, the existing antenatal care (ANC) guidelines are not properly implemented. Hence due to lack of screening and management many women stay uninformed and at risk of adverse outcomes [20]. Literature on nutrition during pregnancy and the postpartum period highlights that Tanzanian women’s diets are often high in calories and predominantly carbohydrate based, with studies showing that 81% of pregnant women exceed recommended carbohydrate intake levels, while 28% have protein intake below recommendations, particularly among those who are overweight or obese [21, 22].

Evidence suggests this high carbohydrate patterns persist postpartum, potentially worsening glycemic control in the absence of targeted guidance. Pregnancy and postpartum offer key healthcare opportunities with eight ANC visits and postnatal care up to six weeks, yet GDM screening is inconsistent, with only 30% of urban women tested.[23, 24]. Obesity among women of reproductive age, is reported at 54% in urban Tanzanian settings, further amplifying the risk of undetected hyperglycemia and GDM [25]. Without intervention, 38% of women with GDM may develop T2DM within 12 weeks postpartum [26]. Current ANC guidelines in Tanzania emphasize nutritional interventions during pregnancy but has not addressed postpartum nutrition for glycemic control, and postpartum follow-up is not routinely practiced [20]. This gap combined with increasing obesity and sedentary lifestyles, emphasize the urgent need for targeted postpartum screening and nutritional interventions to mitigate T2DM risk and improve maternal health outcomes.

This study evaluated postpartum hyperglycemia in women attending postnatal clinics between 6 weeks and 6 months postpartum, using random plasma glucose (RBG ≥7.8 mmol/L or ≥140mg/dL) as the diagnostic criterion for non-pregnant women [27]. By investigating the prevalence of postpartum hyperglycemia and associated nutritional factors in order to fill the knowledge gap in Tanzania’s postpartum care and provide evidence to enhance nutrition counseling and antenatal services and reduce the burden of T2DM.

## Materials and Methods

### Study design and area

We employed an analytical cross-sectional study design to determine the status of postpartum glucose control and assess its association with nutrition and lifestyle factors among women attending postnatal clinics.

The Tanzania health system include national level hospitals, zonal referral hospitals, reginal referral hospitals, district hospitals, health centers, and dispensaries. The majority of reproductive and child health clinics (RCH) attendees are conducted at the health center level [28, 29]. This study was conducted in three public health centers (HC) including Sinza HC, in Ubungo district, Kinyerezi HC in Ilala district and Magomeni HC from Kinondoni district in Dar es salaam, Tanzania. Data on postpartum blood glucose control, physical activity and diet was collected quantitatively using a structured questionnaire from 28^th^ April 2025 to 6^th^ June 2025.

### Study population and sampling

We recruited postpartum women aged 15 to 49 years without a documented Type 2 diabetes mellitus history, attending postnatal clinics at three selected health centers who were six weeks to six months post childbirth, and provided written informed consent. Postpartum women who were ill and under management with known history and/or pre-existing type 1or type 2 diabetes mellitus were excluded.

About 428 postpartum women were approached to participate in the study. However, 36 women (8.4%), refused to participate. The sample size was obtained from the formula by Cochran for infinite unknown population for a prevalence study [30]. The proportion of postpartum hyperglycaemia was set at 50%, with 5% margin of error, 10% non-response rate and 95% confidence interval.

A multistage stratified sampling method was used to determine the three health centers with postnatal (RCH) clinics from the five districts. Postpartum women were randomly selected to participate in the study. At the facility, we collected clinic cards and used a lottery method to select eligible women.

### Variables and measurements

Postpartum hyperglycemia was the dependent variable defined as random blood glucose level (RBG) ≥7.8 mmol/L (≥140mg/dL) according to the American Diabetes Association (ADA) regardless of the time from the last meal [27]. Capillary blood sample was collected for random blood glucose measurements using standardized Glucometer machine (SD Gluco-Navii^®^ GDH). The laboratory research assistant from each HC used aseptic technique to draw 0.5µl of capillary blood sample for measurement using the pricker and test strips. The machines were calibrated by using the control test strip. Duplicate measures were taken for women with hyperglycemia. The disposal of the used strips and prickers was done using the safety box at the facility.

Postpartum Hyperglycemia was categorized as Normal <7.8mmol/L and Hyperglycemia (≥7.8mmol/L). All women with elevated random plasma glucose above the stated range were considered hyperglycemic and were referred to continue with hospital protocol of care.

The independent variables were nutrition status (BMI), diet and physical activity. Nutrition status was assessed using anthropometric measurements. Weight was measured by a mechanical adult beam balance scale with 50g precision, measured to the nearest 0.1kg (Detecto 448 beam balance scale). Height was measured using a stadiometer, without shoes and recorded to the nearest 0.1cm (Detecto 448 beam balance scale). Body mass index was calculated to determine nutrition status as normal, underweight or overweight and obese.

Dietary consumption was determined using a validated Food frequency questionnaire (FFQ) with 20 foods groups commonly consumed in Dar es salaam. The frequency of intake from every food group was categorized as “daily”, “weekly”, “monthly” and “never” if the participant didn’t consume the food. The FFQ was adopted from the validated Africa/Harvard School of Public Health Partnership for Cohort Research and Training (PaCT) questionnaire [31].

Data on physical activity level was collected using the 7-day recall WHO Global physical activity questionnaire adopted and modified for postpartum women in Tanzania. The questionnaire was administered and physical activity was classified as Low physical activity classified as <600 MET and Moderate physical activity ≥ 600 MET. Physical activity was assessed on three domains, activity at work, travel to and from places and recreational activities. Data on other covariates was collected using administered questionnaire. Covariates for the study included Sociodemographic characteristics including maternal age as one’s birthdate, age at conception and marital status (married or not married), Education level and family size. Socioeconomic characteristics including occupation status (employed/unemployed or self-employed) and income level as low, medium and high. Other factors were blood pressure, history of gestational diabetes, breastfeeding, infant birth weight, mode of delivery and history of macrosomia.

### Data collection and quality assurance

Two research assistants were recruited to assist with data collection. Where one was an assistant nursing officer and another was a laboratory technician. They were trained and oriented on the research proposal, objectives, data collection and ethical considerations. They went through a one week in depth training program on the data collection tools, the Food Frequency Questionnaire, Global physical activity questionnaire, structured questionnaire on covariates and ethical issues. Data collection was done sequentially on the three health facilities. The Swahili version of the questionnaires was uploaded in the Kobo toolbox and tested on the pilot population at Kimara Health center and Kawe Health center which were not part of the main study. The collected data were cleaned, processed, and securely stored in the personal password-protected computer. To ensure data quality, each day, data was inspected for inconsistency and incompleteness and further cleaning was done during data analysis.

### Data management and analysis

The data was analyzed using Stata version 17 (Stata Corp, College Station, TX, USA). Descriptive statistics was used to summarize the data on dependent and independent variables including frequencies for categorical variables, proportions, means, standard deviations, median and range for numerical variables depending on the distribution. The categorical variables were compared between women with postpartum hyperglycemia and those with normal blood glucose using Pearsońs chi square test. Modified poisson regression analysis was used to identify the factors with statistically significant association between the outcome variable and the independent variables. Univariable analysis was done to determine individual association. Also, factors with p<0.2 and those of scientific significance were included in the multivariable poisson regression model. To determine independent association after controlling for confounders. Statistical significance was set at p<0.05. Data on dietary consumption was analyzed using Principal component analysis (PCA). And data on Physical activity was analyzed in excel where metabolic equivalent of tasks were calculated and categorized according to WHO [32].

### Ethical consideration

Ethical clearance was obtained from the Muhimbili University of Health and Allied Sciences (MUHAS) Institutional Review Board (IRB) reference No. DA.282/298/01.C/2587. Written informed consent was obtained from the participating women.

## Results

### Socio-demographic characteristics of participants

The study enrolled 392 postpartum women. The mean age was 31.5 years (SD ±7.54), with the sample that was normally distributed across age groups, where below 35 years was (69.1%) while (30.8%) over 35 years. Participants were recruited from three health facilities, with Sinza HC contributing (48.0%), followed by Magomeni HC (28.3%) and Kinyerezi HC (23.7%). Most participants were not married (67.6%), and the majority had secondary (44.6%) or primary (38.0%) education, while only 17.6% had above high school education. Over half (52.6%) were self-employed, 34.4% were unemployed, and 13.0% were employed. Economic status varied, with 52.3% earning less than 500,000 Tshs monthly, 29.8% earning 500,000–1,000,000 Tshs, and 17.9% earning over 1,000,000.

### Prevalence of Postpartum Hyperglycemia

The prevalence of postpartum hyperglycemia (≥7.8 mmol/L) was 18.6% (n=73), while 81.4%, (n=319) of the study participants had normal random blood glucose (RBG) levels (<7.8 mmol/L).

### Dietary consumption of the study participants

Table below (table 1) summarizes the median and interquartile range (IQR) of the five dominant principal components consumed by study participants that were renamed into dietary patterns based on the dominant foods from (PCA). The median reflects the central tendency of adherence to each dietary pattern, while the Interquartile range reflects variability in adherence among participants. The most dominant principal components (patterns) among the study participants were Pattern 1 (Refined carbohydrate) stiff porridge 0.469, white bread 0.527 and sweet potatoes were dominant 0.312, and Pattern 2 (Legumes and mixed diet) with more legume consumption such as beans 0.393, Pattern 3 (Animal protein) with beef 0.327, Pattern 4 (Sweetened beverages) 0.140, and Pattern 5 (Fats) 0.412.

**Table 1:**
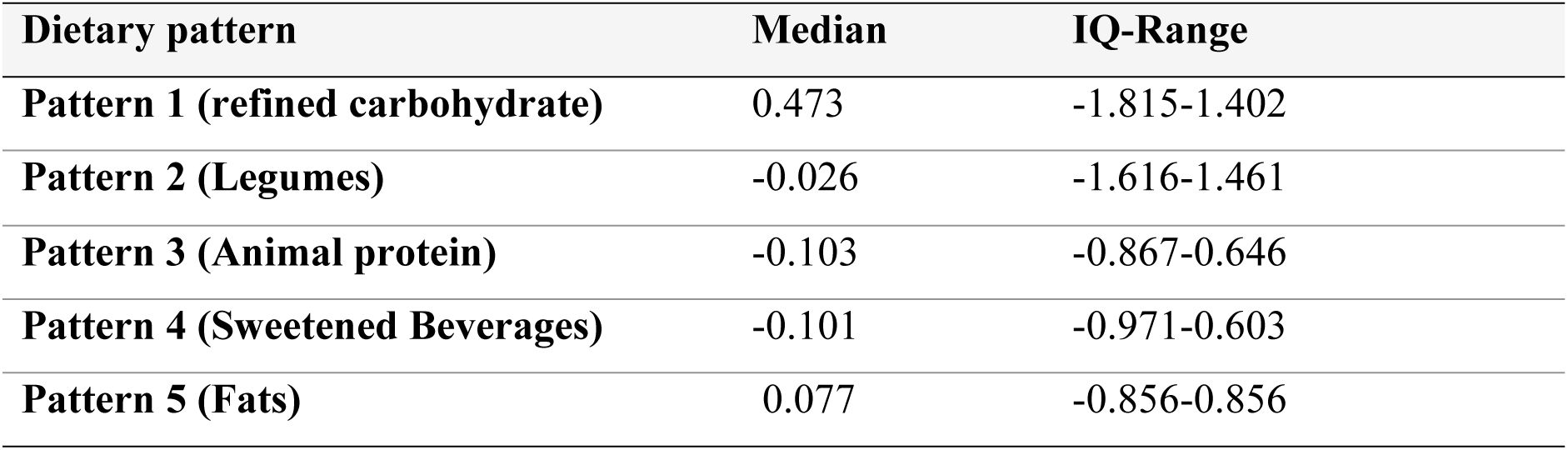
Dietary pattern of the study participants.

Among the five patterns, refined carbohydrate pattern, showed a significant positive association with postpartum hyperglycemia, while the legume-rich pattern showed a protective effect towards postpartum hyperglycemia. Pattern 1 (refined carbohydrate), includes high intake of stiff porridge, bread, and refined grains. The positive median is corresponding to positive loadings showing that many participants followed this pattern which was associated with increased prevalence of postpartum hyperglycemia (APR = 1.54, p = 0.022). Pattern 2 (Legumes), featured beans and green vegetables with moderate distribution. This pattern was shown to be Protective against hyperglycemia (APR = 0.69, p = 0.001). this indicated a diverse diet including plant-based proteins and vegetables that may help reduce glucose abnormalities postpartum.

Dietary pattern three has a negative median implying low to moderate intake of animal-based protein foods among most participants, while pattern four participants consumed sweetened beverages less frequently, though some variation exists. The median for dietary pattern five indicates moderate adherence to fat-rich foods (e.g., oils, butter), with a fairly balanced intake among participants.

### Distribution of participants by potential risk factors for postpartum hyperglycemia

With regards to age, the difference observed between older women and younger women was statistically significant. Hyperglycemia was more prevalent on older women 45.4% among those over 35 years as compared to eighteen cases among those below 35 years 6.6%. Also, education was a significant factor where women with primary education or lower had a higher prevalence of postpartum hyperglycemia (25.0%) as compared to those with secondary school education (17.1%) or higher (8.7%). Moreover, Socio-economic status and occupation also observed a significant difference between women with postpartum hyperglycemia and those without with 32.6% of those unemployed having hyperglycemia. Women with higher BMI 50% of obese and 39.4% of overweight participants had hyperglycemia, compared to only 2% of those with normal BMI. Physical activity, history of hypertension during pregnancy, higher parity, marital status and breastfeeding length observed significance difference among women with postpartum hyperglycemia and normal glycemic women.

### Factors associated with postpartum hyperglycemia

In univariable analysis, significant risk factors included BMI overweight and obese with higher prevalence, low physical activity, dietary patterns Pattern 1 increased prevalence, (CPR=2.52, p<0.001) Patterns 2, 3 and 4 were protective, age (<35 years protective, CPR=0.03, p<0.001), socio-economic status middle and higher income was protective, parity (lower parity protective, p=0.021), unemployment (CPR=2.07, p=0.036), History of hypertension during pregnancy (CPR=1.83, p=0.004), unmarried status (CPR=2.52, p=0.007), and normal birthweight protective, (CPR=0.33, p=0.00). In multivariable analysis, overweight and obesity (higher BMI) and diet was identified as key modifiable risk factors for postpartum hyperglycemia, (APR = 8.1, 11.6), p <0.001) and (APR=1.54, 0.69, p 0.022, 0.001) respectively.

Higher (BMI) was strongly associated with increased prevalence of postpartum hyperglycemia, with participants having a higher BMI showing a significantly higher prevalence of hyperglycemia compared to those who were normal (adjusted prevalence ratio (APR=8.1, 11.6, p<0.001). Additionally, diet played a significant role: Pattern 1 (Refined carbohydrate) was associated with an increased risk of postpartum hyperglycemia (APR=1.54, 95% CI: 1.13–2.26, p=0.022), while Pattern 2 (legumes and vegetables) was protective, reducing the risk (APR=0.69, 95% CI: 0.55– 0.88, p=0.001), (Refer table 3). While physical activity levels expressed as low physical activity and moderate physical activity in MET per minutes showed no association with postpartum hyperglycemia in the multivariable model (APR= 1.02, p=0.927).

**Table 2:**
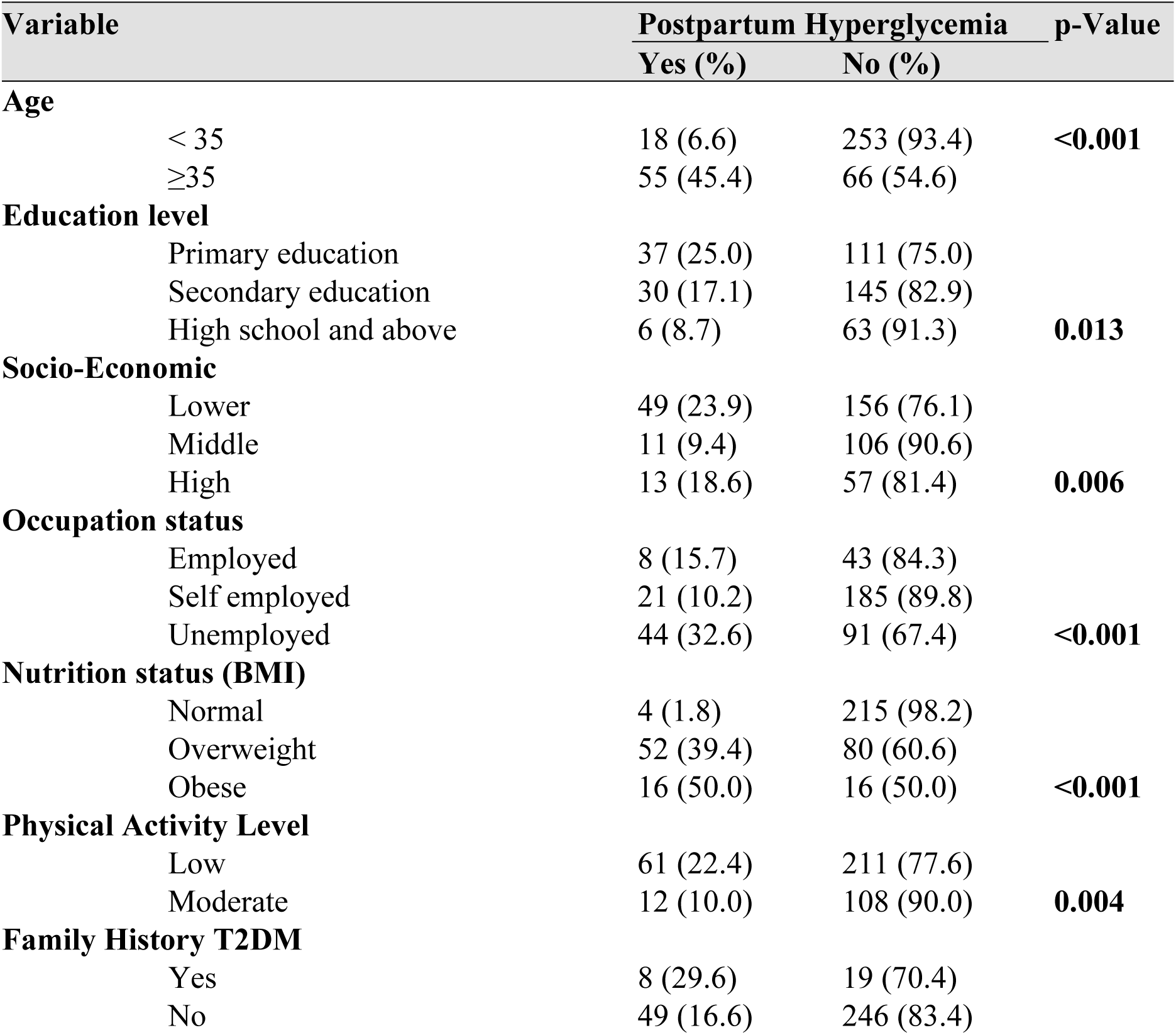
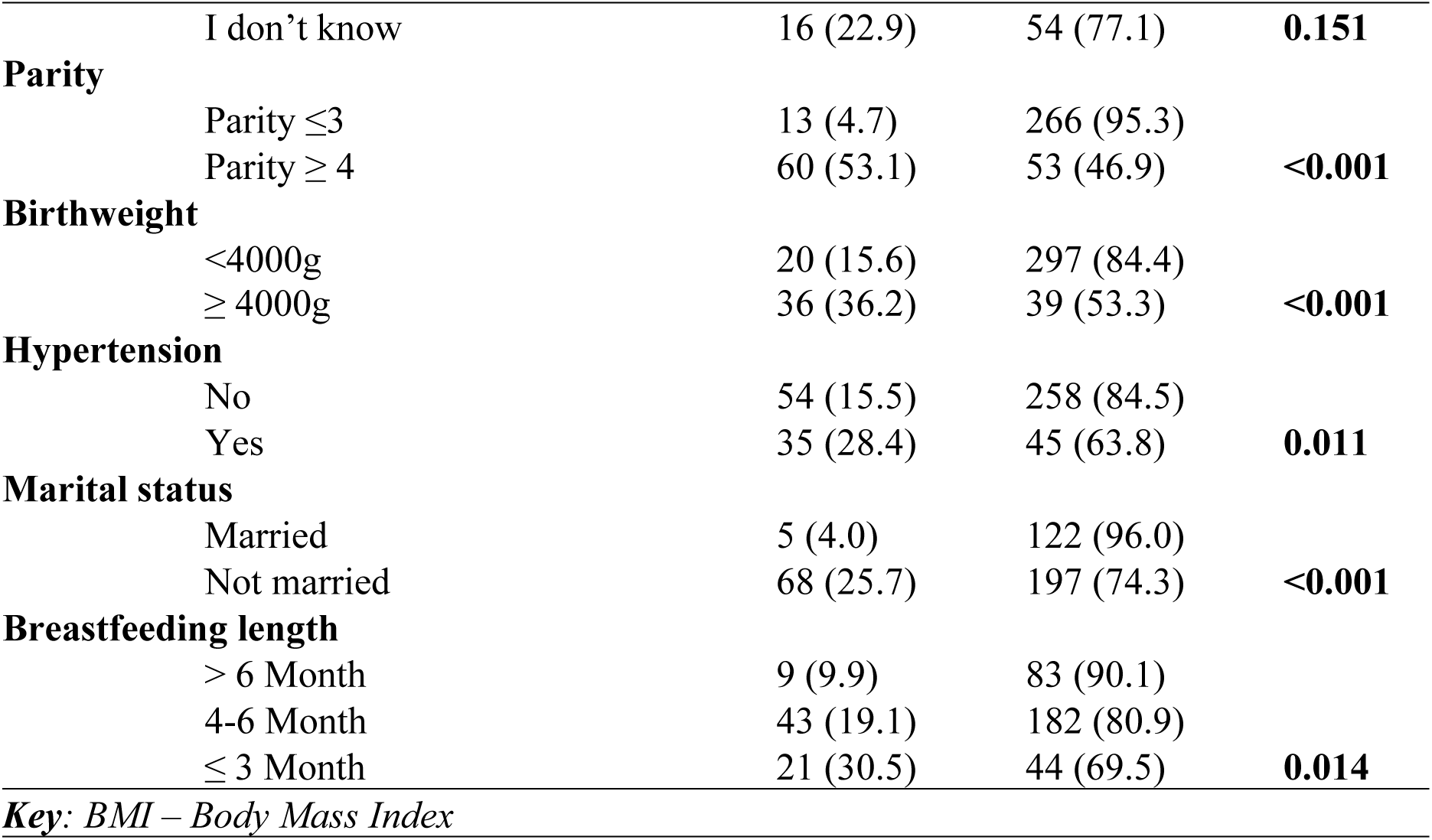
Distribution of participants with potential factors for postpartum hyperglycemia (n=392).

**Table 3:**
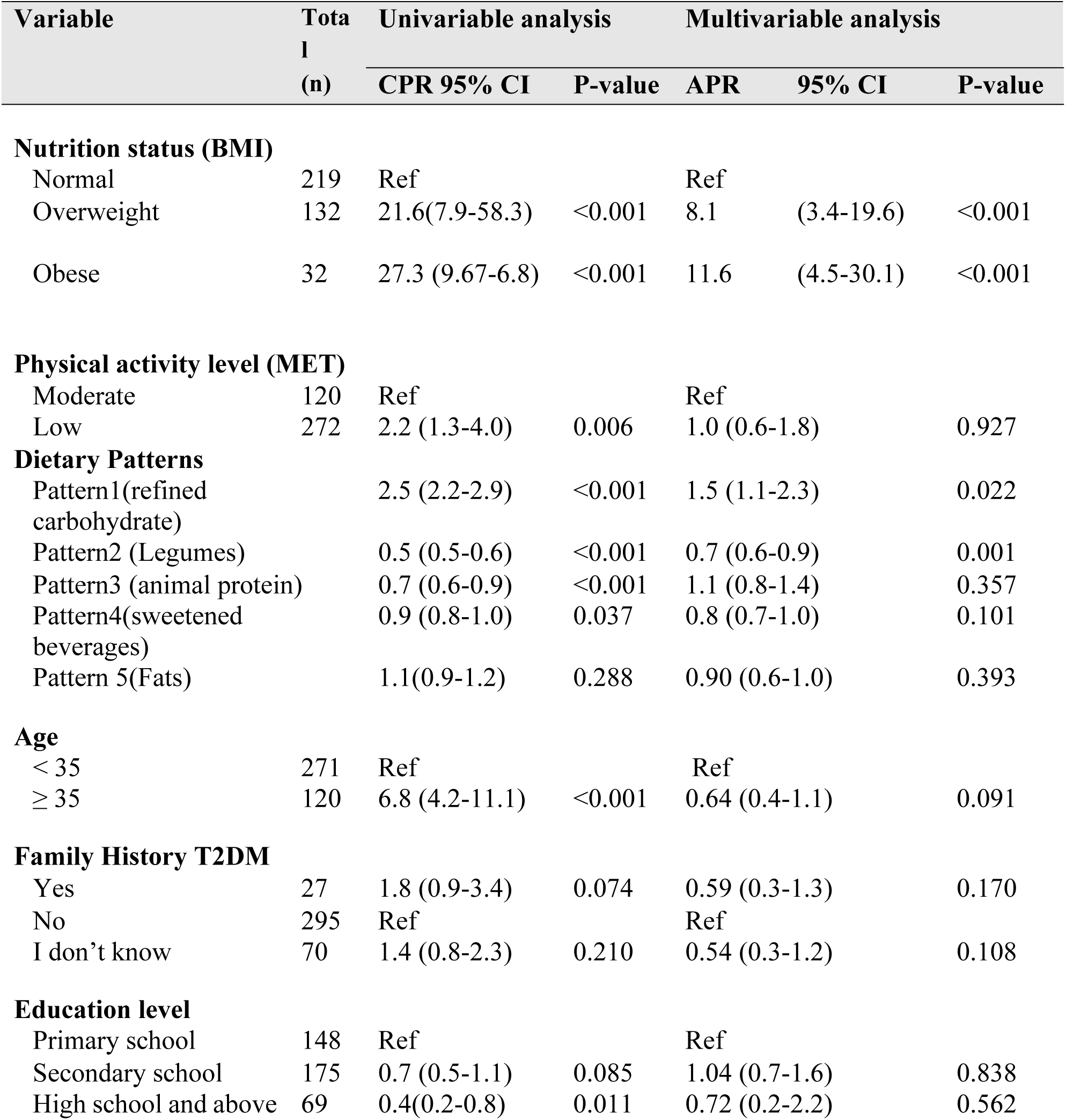
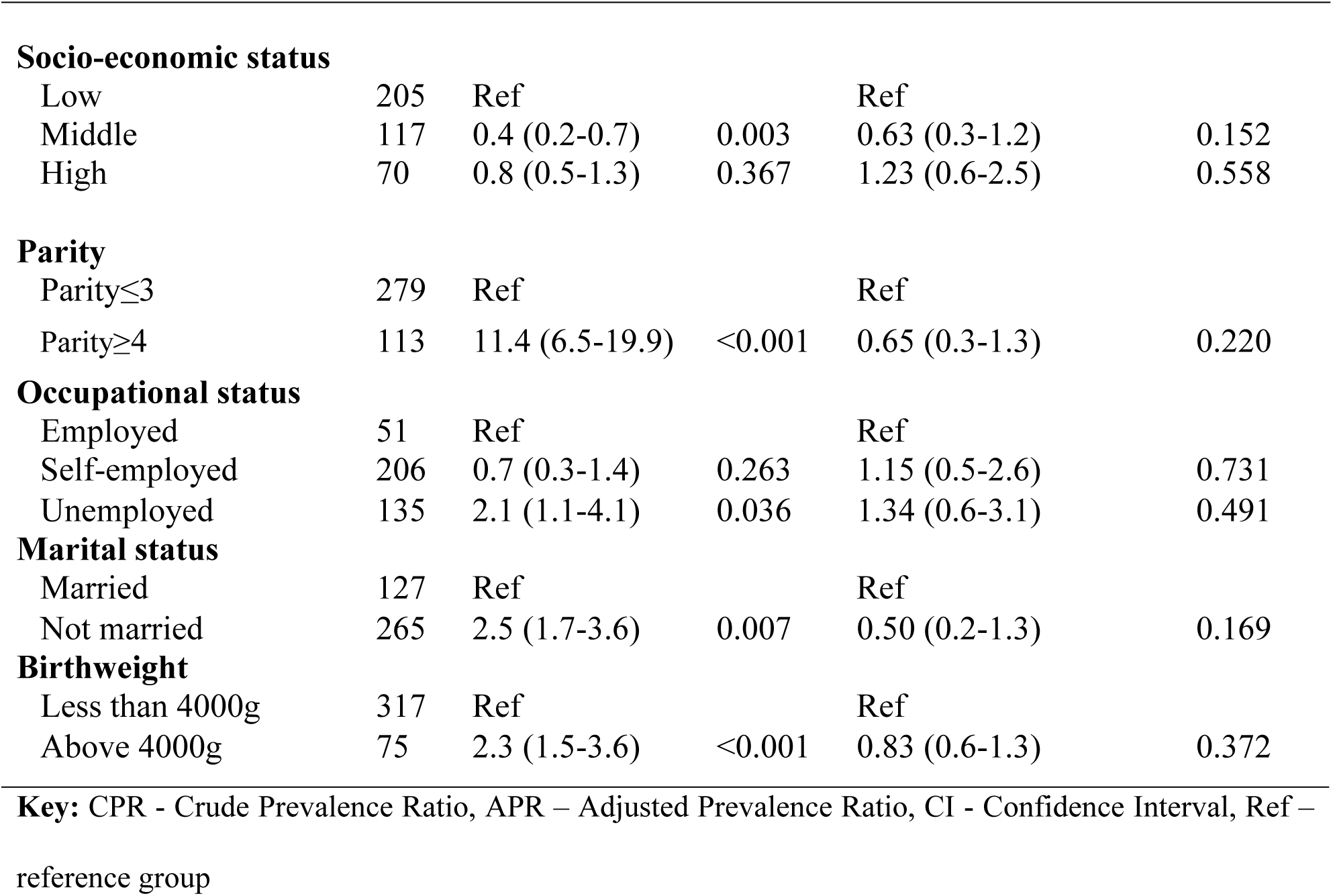
Univariable and multivariable analysis of factors associated with postpartum hyperglycemia.

## Discussion

Postpartum hyperglycemia was prevalent among 18.6% of women attending postnatal clinics in Dar es Salaam. Further, hyperglycemia in this vulnerable population was associated with overweight, obesity, and consumption of refined carbohydrate; while protein and legumes were protective factors. This prevalence underscores a significant public health challenge in urban Tanzanian settings, where metabolic disorders are rising due to increasing obesity and sedentary lifestyles [21].

The unprecedented burden of postpartum hyperglycemia in our study aligns with regional studies in Ghana [33], but slightly lower than other contexts in the continent [34]. It should be noted that, persistent hyperglycemia is a risk factor for progressing to type 2 diabetes mellitus (T2DM), also noted in South Africa [35], and other contexts. In a wake of heightened burden diabetes in relatively younger populations in sub–Saharan Africa, and particularly among women, it is imperative to address and mitigate the determinants of postpartum hyperglycemia. Such determinants as found in this study are also increasing owing to economic, demographic, and nutritional transitions [21].

Overweight and obesity were independently associated with postpartum hyperglycemia in this study. Evidence presented in this study is consistent with studies from South Africa, Brazil, and Spain [35–37]. Overweight and obesity resulted excess body fat is linked with insulin resistance and impaired glucose metabolism. Metabolic dysfunction owing to reducing insulin sensitivity is a well-documented pathway to hyperglycemia and chronic type 2 diabetes, especially in women with prior gestational diabetes [38, 39]. With the growing burden of overweight and obesity in the country, particularly among women of reproductive age, pre-pregnant excessive weight and fat accumulation will in turn increase the T2DM unless addressed.

Tanzania is undergoing a nutritional transition with increasing energy and caloric intake. About 81% of Tanzanian pregnant women exceed recommended carbohydrate intake, often from refined sources like white rice and maize, which are rapidly digested, causing blood glucose spikes and stressing pancreatic function [21, 22]. Conversely, diets rich in legumes, lean proteins, and vegetables was protective, aligning with evidence that low glycemic index foods improve insulin sensitivity [36]. Evidence from this study indicate a clear association between certain dietary patterns with postpartum hyperglycemia. Tanzania’s Food Based Dietary Guidelines recommend a balanced diet with increased protein for lactating women, but socioeconomic barriers, lack of awareness, and cultural preferences for carbohydrate heavy meals hinder implementation [40].

Dietary pattern, combined with postpartum hormonal changes that impair glucose metabolism, increase T2DM risk [41]. The guidelines, however, lack of specific postpartum nutritional recommendations represents a critical gap in addressing glycemic control [20].

Physical activity levels, measured in METs per minutes, showed no significant association with postpartum hyperglycemia in the multivariable model, suggesting that activity alone may not mitigate hyperglycemia without addressing BMI or diet. This null effect may stem from misclassification, short postpartum duration, or social desirability bias in self-reported activity [42]. However, physical activity remains a modifiable factor for glucose control and should be promoted alongside dietary interventions [19]

Age over 35 years was a significant risk factor, with older women showing higher hyperglycemia prevalence (48.3%) compared to younger women (6.6%), presumed to be due to reduced pancreatic beta-cell function and increased fat mass, which impair insulin production and sensitivity [16, 38, 41]. These findings underscore the need for age specific interventions, particularly as postpartum hormonal fluctuations exacerbate glucose intolerance following pregnancy [41].

Although this cross-sectional study is subject to limitations such as inability to determine causality, it used standard and validated tools to collect data on factors associated with postpartum hyperglycemia. Moreover, multivariable modified poisson regression analysis was used to account for potential confounding variables such as age, parity, and socioeconomic status strengthens the internal validity of the results. Also, the study was multicentered which enhance generalizability. However, we encourage future studies to include multiple measurements such as HbA1c which could give a more detailed data on long-term glucose control.

## Conclusion

Approximately one in five postpartum women in Dar es Salaam attending postnatal clinics had hyperglycemia, indicating a significant public health concern. This high prevalence is driven by overweight and obesity and high consumption of refined carbohydrates, which can exacerbated insulin resistance and glucose dysregulation, particularly in older women. To mitigate type 2 diabetes risk, attention should be put to prioritizing nutrition education and promotion of healthy lifestyle during peripartum period to ensure optimal glucose control in the postpartum period.

## Data Availability

All relevant data are within the manuscript and its Supporting Information files.

## Acknowledgement

The author wishes to acknowledge dedicated academic staff at MUHAS from the School of Public Health, Department of Community health and College of clinical medicine, the women who participated fully in the study, PORALG and dedicated staffs from the health centers in Dar es salaam for their guidance, expertise, mentorship and support during conceptualization and implementation of the study. The author of this Master’s thesis was supported by GROWNUT-II project, a NORAD-II funded partnership project and the EPRENUT program a NORPART funded capacity building for nutritional epidemiology at MUHAS.

## Reference

[1] Practice C. Diagnostic criteria and classification of hyperglycaemia first detected in pregnancy: a World Health Organization Guideline. Diabetes Res Clin Pract 2014; 103: 341–363.

[2] Coetzee A, Sadhai N, Mason D, et al. Evidence to support the classification of hyperglycemia first detected in pregnancy to predict diabetes 6–12 weeks postpartum: A single center cohort study. Diabetes Res Clin Pract 2020; 169: 108421.

[3] Kramer CK, Campbell S, Retnakaran R. Gestational diabetes and the risk of cardiovascular disease in women: a systematic review and meta-analysis. Diabetologia 2019; 62: 905–914.

[4] Sweeting A, Hannah W, Backman H, et al. Series Gestational Diabetes 2 Epidemiology and management of gestational diabetes. 2024; 6736: 1–18.

[5] Hivert M, Backman H, Benhalima K, et al. Series Gestational Diabetes 1 Pathophysiology from preconception, during pregnancy, and beyond. Lancet 2024; 6736: 1–17.

[6] Norris SA, Zarowsky C, Murphy K, et al. Integrated health system intervention aimed at reducing type 2 diabetes risk in women after gestational diabetes in South Africa (IINDIAGO): A randomised controlled trial protocol. BMJ Open 2024; 14: 1–11.

[7] Coetzee A, Hall DR, van de Vyver M, et al. Early postpartum HbA1c after hyperglycemia first detected in pregnancy—Imperfect but not without value. PLoS One; 18. Epub ahead of print 2023. DOI: 10.1371/journal.pone.0282446.

[8] Utz B, De Brouwere V. ‘Why screen if we cannot follow-up and manage?’ Challenges for gestational diabetes screening and management in low and lower-middle income countries: Results of a cross-sectional survey. BMC Pregnancy Childbirth; 16. Epub ahead of print 2016. DOI: 10.1186/s12884-016-1143-1.

[9] Mwanri AW, Kinabo J, Ramaiya K, et al. Prevalence of gestational diabetes mellitus in urban and rural Tanzania. Diabetes Res Clin Pract 2013; 103: 71–78.

[10] Grunnet LG, Hjort L, Minja DT, et al. High Prevalence of Gestational Diabetes Mellitus in Rural Tanzania — Diagnosis Mainly Based on Fasting Blood Glucose from Oral Glucose Tolerance Test.

[11] Ruhembe CC, Mosha TCE, Nyaruhucha CNM. Diabete 2 Mellitis. Tanzania J Heal Res Vo 2014; 16: 1–11.

[12] Yuste Gómez A, del Ramos Álvarez M P, Bartha JL. Influence of Diet and Lifestyle on the Development of Gestational Diabetes Mellitus and on Perinatal Results. Nutrients 2022; 14: 1–14.

[13] Dolatkhah N, Hajifaraji M, Shakouri SK. Nutrition therapy in managing pregnant women with gestational diabetes mellitus: A Literature Review. J Fam Reprod Heal 2018; 12: 57– 72.

[14] Msollo SS, Martin HD, Mwanri AW, et al. Simple method for identification of women at risk of gestational diabetes mellitus in Arusha urban, Tanzania. BMC Pregnancy Childbirth 2022; 22: 1–11.

[15] Dludla P V, Mabhida SE, Ziqubu K, et al. Pancreatic β-cell dysfunction in type 2 diabetes: Implications of inflammation and oxidative stress. World J Diabetes 2023; 14: 130–146.

[16] Mdoe MB, Kibusi SM, Munyogwa MJ, et al. Prevalence and predictors of gestational diabetes mellitus among pregnant women attending antenatal clinic in Dodoma region, Tanzania: an analytical cross-- sectional study. 2021; 69–79.

[17] Mcdonald SM, May LE, Hinkle SN, et al. Maternal Moderate-to-Vigorous Physical Activity before and during Pregnancy and Maternal Glucose Tolerance: Does Timing Matter? Med Sci Sports Exerc 2021; 53: 2520–2527.

[18] Xie W, Zhang L, Cheng J, et al. Physical activity during pregnancy and the risk of gestational diabetes mellitus: a systematic review and dose–response meta-analysis. BMC Public Health; 24. Epub ahead of print 2024. DOI: 10.1186/s12889-024-18131-7.

[19] Yaping X, Huifen Z, Meijing Z, et al. Effects of Moderate-Intensity Aerobic Exercise on Blood Glucose Levels and Pregnancy Outcomes in Patients With Gestational Diabetes Mellitus: A Randomized Controlled Trial. Diabetes Ther 2021; 12: 2585–2598.

[20] United THE, Of R, Health MOF, et al. Antenatal care guidelines, MoH.

[21] Killel E, Mchau G, Mbilikila H, et al. Dietary intake and associated risk factors among pregnant women in Mbeya, Tanzania. PLOS Glob Public Heal 2024; 4: e0002529.

[22] Ruhighira JJ, Dionis I, Munyogwa M, et al. High-calorie low-protein dietary pattern among overweight and obese pregnant women in Tanzania. Clin Nutr Open Sci 2025; 60: 89–100.

[23] Mukuve A, Noorani M, Sendagire I, et al. Magnitude of screening for gestational diabetes mellitus in an urban setting in Tanzania; A cross-sectional analytic study. BMC Pregnancy Childbirth 2020; 20: 1–8.

[24] Kikula A, Sirili N, Ramaiya K, et al. Optimizing screening practice for gestational diabetes mellitus in primary healthcare facilities in Tanzania: research protocol. Reprod Health 2024; 21: 193.

[25] Ministry of Health T. Tanzania Demographic Health Survey-MIS 2022. TDHS-MIS 2022.

[26] Msollo SS, Mwanri AW. Postpartum hyperglycemia and pregnancy outcomes among women in Arusha Region, Tanzania. Tanzan J Health Res 2022; 23: 1–12.

[27] Tests D, Diabetes FOR. 2. Classification and diagnosis of diabetes. Diabetes Care 2016; 39: S13–S22.

[28] Kwesigabo G, Mwangu MA, Kakoko DC, et al. Tanzania’s health system and workforce crisis. J Public Health Policy 2012; 33: S35–S44.

[29] MoH. United Republic of Tanzania Ministry of Health, Community Development, Gender, Elderly and Children Health Sector Strategic Plan Leaving No One Behind, https://www.moh.go.tz/storage/app/uploads/public/672/630/145/6726301451e92438381140.pdf (2021).

[30] Bartlett II JE, Kotrlik JW, Higgins CC. Determing appropriate sample size in survey research. Inf Technol Learn Perform J 2001; 19: 43–50.

[31] Malindisa E, Dika H, Rehman AM, et al. Dietary patterns and diabetes mellitus among people living with and without HIV: a cross-sectional study in Tanzania. Front Nutr 2023; 10: 1–11.

[32] WHO. Global Physical Activity Questionnaire.

[33] Agbozo F, Abubakari A, Zotor F, et al. Gestational diabetes mellitus per different diagnostic criteria, risk factors, obstetric outcomes and postpartum glycemia: A prospective study in Ghana. Clin Pract 2021; 11: 257–271.

[34] Coetzee A, Hall DR, Conradie M. Hyperglycemia First Detected in Pregnancy in South Africa: Facts, Gaps, and Opportunities. Front Clin Diabetes Healthc 2022; 3: 1–18.

[35] Chivese T, Norris SA, Levitt NS. Progression to type 2 diabetes mellitus and associated risk factors after hyperglycemia first detected in pregnancy: A cross-sectional study in Cape Town, South Africa. PLoS Med 2019; 16: 1–18.

[36] Dias LM, Schmidt MI, Vigo Á, et al. Dietary Patterns in Pregnancy and the Postpartum Period and the Relationship with Maternal Weight up to One Year after Pregnancy Complicated by Gestational Diabetes. Nutrients 2023; 15: 1–14.

[37] Li LJ, Huang L, Tobias DK, et al. Gestational Diabetes Mellitus Among Asians – A Systematic Review From a Population Health Perspective. Front Endocrinol (Lausanne); 13. Epub ahead of print 2022. DOI: 10.3389/fendo.2022.840331.

[38] Civantos S, Durán M, Flández B, et al. Predictors of postpartum diabetes mellitus in patients with gestational diabetes. Endocrinol Diabetes y Nutr 2019; 66: 83–89.

[39] Miao Z, Wu H, Ren L, et al. Long-Term Postpartum Outcomes of Insulin Resistance and β-cell Function in Women with Previous Gestational Diabetes Mellitus. Int J Endocrinol; 2020. Epub ahead of print 2020. DOI: 10.1155/2020/7417356.

40. MoH. Ministry of Health, Community Development, Gender, Elderly and Children (MoHCDGEC). Tanzania Food-Based Diet Guidel, https://www.fao.org/nutrition/education/food-dietary-guidelines/regions/tanzania/en/ (2011).

[41] Rassie K, Giri R, Joham AE, et al. Human Placental Lactogen in Relation to Maternal Metabolic Health and Fetal Outcomes: A Systematic Review and Meta-Analysis. Int J Mol Sci; 23. Epub ahead of print 2022. DOI: 10.3390/ijms232415621.

[42] Dipla K, Zafeiridis A, Mintziori G, et al. Exercise as a Therapeutic Intervention in Gestational Diabetes Mellitus. Endocrines 2021; 2: 65–78.

